# A Multi-layer Perceptron Neural Network for Predicting the Osteoporosis in Women Using Physical Activity Factors

**DOI:** 10.1101/2023.03.06.23286695

**Authors:** Jun-hee Kim

**Affiliations:** Yeonsedae-gil, Maeji-ri, Heungeop-myeon, Wonju-si, Gangwon-do, 26493, Laboratory of KEMA AI Research (KAIR), Department of Physical Therapy, College of Software and Digital Healthcare Convergence, Yonsei University, Wonju, South Korea

**Author notes:** **Corresponding author:** Jun-hee Kim (JH Kim), PhD, PT, Address: 1, / /.

**Keywords:** Osteoporosis, predictive model, multi-layer perceptron, deep learning, physical activity

## Abstract

**Introduction:** Osteoporosis (OP) is a bone disease caused by a decrease in bone mineral density (BMD). OP is common in women because BMD gradually decreases after age 35. OP due to decreased BMD is highly likely to cause fatal traumatic injuries such as hip fracture. The purpose of this study was developed and evaluated a multi-layer perceptron neural network model that predicts OP using physical characteristics and activity factors of adult women over the age of 35 whose BMD begins to decline.

**Materials and Methods:** Data from KNHANES were used to develop a multi-layer perceptron model for predicting OP. Data preprocessing included variable selection and sample balancing, and LASSO was used for feature selection. The model used 5 hidden layers, dropout and batch normalization and was evaluated using evaluation scores such as accuracy and recall score calculated from a confusion matrix.

**Results:** Models were trained and evaluated to predict OP using selected features including age, quality of life index, weight, grip strength and average working hours per week. The model achieved 76.8% accuracy, 74.5% precision, 80.5% recall, 77.4% F1 score, and 74.8% ROC AUC.

**Conclusion:** A multi-layer perceptron neural network for predicting OP diagnosis using physical characteristics and activity factors in women aged 35 years or older showed relatively good performance. Since the selected variables can be easily measured through surveys, assessment tool, and digital hand dynamometer, this model will be useful for screening elderly women with OP or not in areas with poor medical facilities or difficult access.

## Introduction

Osteoporosis (OP) is a bone disease that occurs when bone mineral density (BMD) and bone mass decrease [1]. In other words, a decrease in BMD below a certain level is regarded as the onset of OP [1]. In women, rapid BMD and skeletal growth occur from menarche, when the secretion of sex steroids, including estrogen, increases [2]. BMD in adult women peaks between the ages of 25 and 35, after which women experience a gradual decline throughout their lives [2]. A decrease in BMD is prominent from menopause, when female hormones decrease, resulting in a higher rate of OP than in men [3, 4]. In particular, estrogen is thought to be important for maintaining BMD, and it has been reported that bone loss may occur as estrogen levels drop during menopause, resulting in a much higher rate of OP [5, 6].

Patients diagnosed with OP have a high risk of fracture due to trauma compared to normal people [7–9]. In particular, the femur, which makes up the hip joint, is a part where fractures commonly occur due to OP, and hip fracture can cause great difficulties in independent activities of daily life, and in severe cases, can be life threatening [10]. The 1-year mortality rate due to hip fracture increases by 2% every year, and in the case of women, the 10-year mortality rate after hip fracture is reported to be 16% [11]. In addition, once a fracture occurs, the risk of a second fracture increases more than twice, and when a second fracture occurs, the patient’s mortality rate is much higher [12]. The mortality within 42 months after a second hip fracture was 57.2% [13]. Because the mortality rate of patients with hip fractures is high, it is important to predict or diagnose OP early to prevent hip fractures from occurring.

BMD is measured using dual energy X-ray absorptiometry (DXA) scans of the spine and hip to diagnose OP [14]. For postmenopausal women, BMD is determined by T-score compared to reference values of race- and sex-matched young adults [14]. However, one of the disadvantages of diagnosing OP through DXA scan is that measurement errors may occur due to surrounding soft tissue [15]. Therefore, recently, studies applying machine learning such as deep learning to predict OP with high performance or to predict OP with a simpler X-ray imaging technique have been actively conducted. Learning hip radiographs and clinical data through deep learning can improve diagnostic performance for OP classification and prediction even without using the DXA scans [16]. Similarly, diagnosis using deep learning based on lumbar radiographs had the potential to screen for OP and osteopenia even without using DXA scans [17]. In addition, The CT image-based deep learning diagnosis method showed higher accuracy and could output higher specificity or sensitivity for OP evaluation [18].

Deep learning models that predict OP using these medical diagnostic devices have been studied, but deep learning models that predict OP according to data that can be acquired without professional knowledge, such as physical characteristics or activity levels, have not been studied. Bone is an organ that continuously undergoes remodeling processes of resorption and formation, and BMD is formed by the balance of these processes [19]. According to the physical stress theory, bone can remain constant in an environment given an appropriate amount of stress [20]. In other words, factors that regulate stress on bones, such as physical activity, are important in controlling BMD [20]. Low activity level was considered a risk factor for OP, and vigorous physical activity such as strength training or aerobic exercise was recommended to prevent OP [21, 22]. However, despite the benefits of moderate loads for BMD, too much activity due to overtraining at elite sports levels can have negative consequences in BMD.[23] Moderate intensity physical activity may help prevent OP, but excessive intensity physical activity may increase the risk of OP [24, 25]. Therefore, data related to physical activity will be able to be trained through deep learning as factors for predicting OP.

The purpose of this study was to create a multi-layer perceptron neural network model that predicts the diagnosis of OP in adult women with a high risk of OP using physical characteristics and physical activity-related factors, and to evaluate the performance of the created multi-layer perceptron deep learning model.

## Materials and Methods

### 1. Data Source

This study used data samples obtained from the 6th to 8th KNHANES (2015-2019), the national statistics of the Korea Disease Control and Prevention Agency. From a total of 39,759 data samples, data were extracted from women aged 35 years or older, the age at which bone density begins to decline. Data samples with missing values were excluded, and 11,929 samples were finally selected. The selected data samples were classified into 9915 data samples of normal subjects and 2014 data samples of subjects diagnosed with OP (Figure 1). This study was approved by the Public Institutional Review Board Designated by Ministry of Health and Welfare (approval number: P01-202303-01-003)

**Figure 1.**
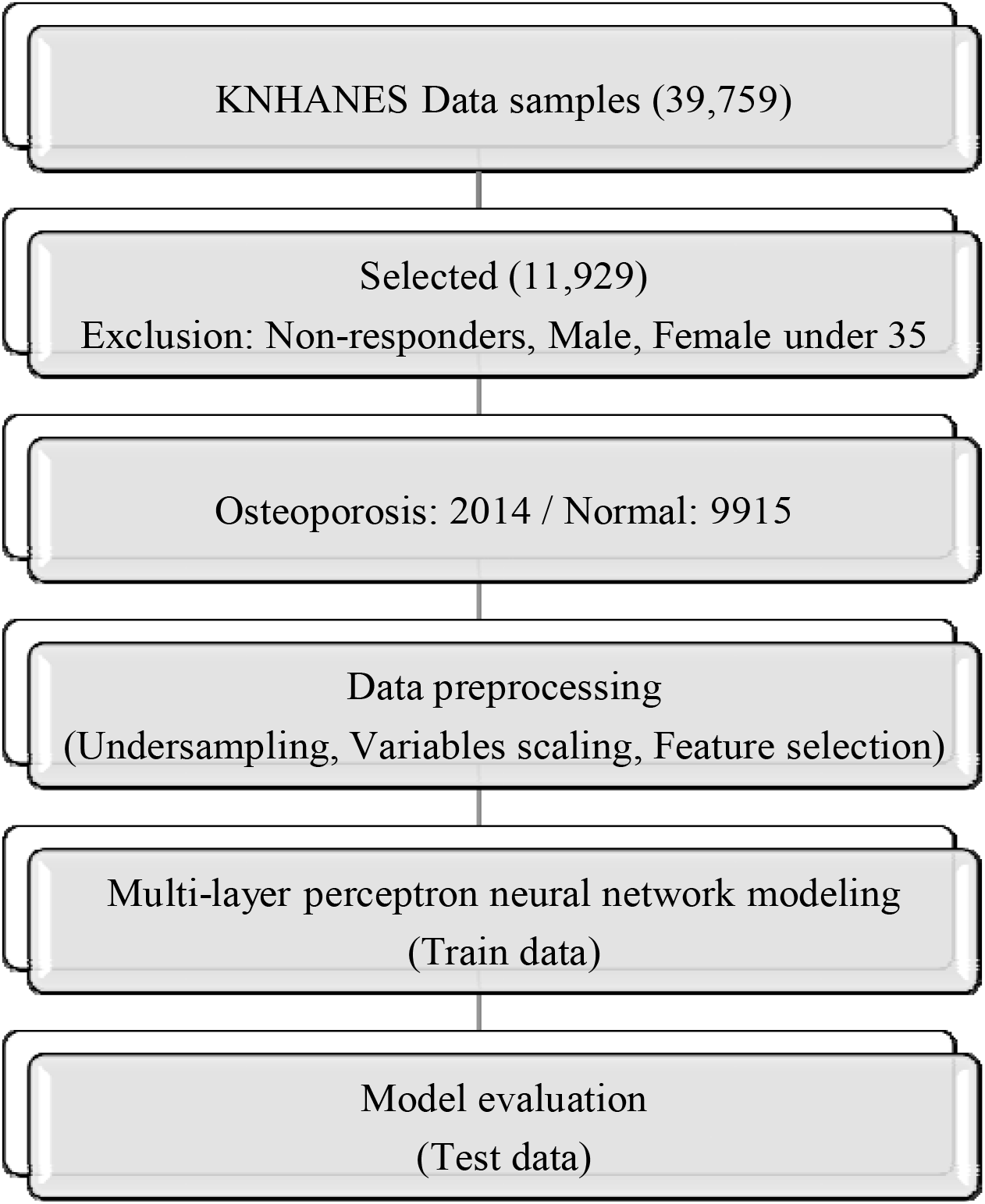
Flowchart of the study.

**Figure 2.**
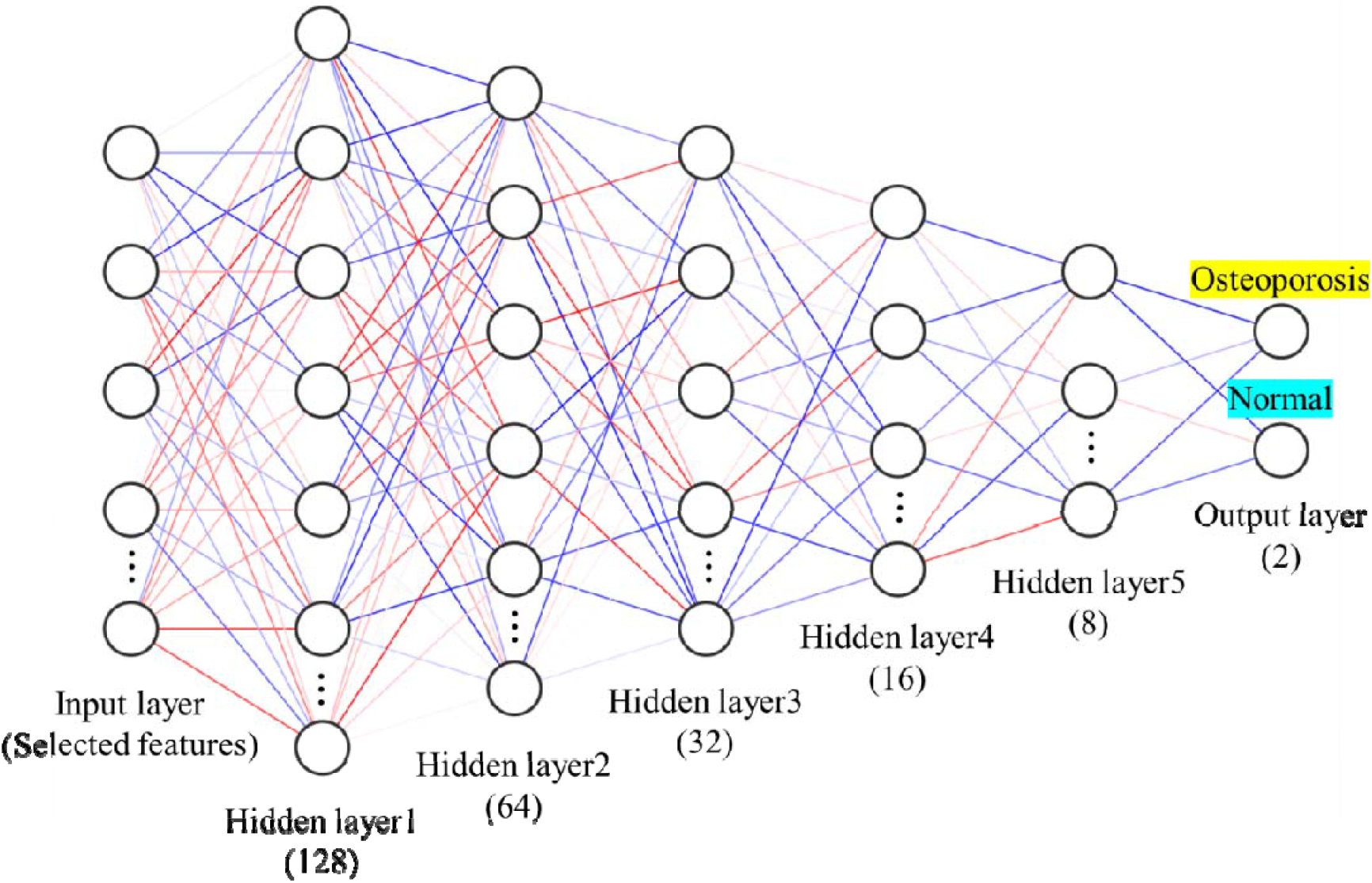
Schematic diagram of multi-layer perceptron neural network.

### 2. Data Preprocessing and Feature Selection

The physical characteristics and activity variables of the subjects were extracted from the data for analysis. Age, weight, height, waist circumference, grip strength, body mass index, quality of life, average working hours per week, walking days per week, and strength training days per week were selected as continuous variables. Activity restriction, occupational activity, work-related moderate-intensity physical activity, work-related moderate-intensity physical activity, leisure-related high-intensity physical activity, leisure-related moderate-intensity physical activity, and weight change were selected as categorical variables (Table 1).

**Table 1.**
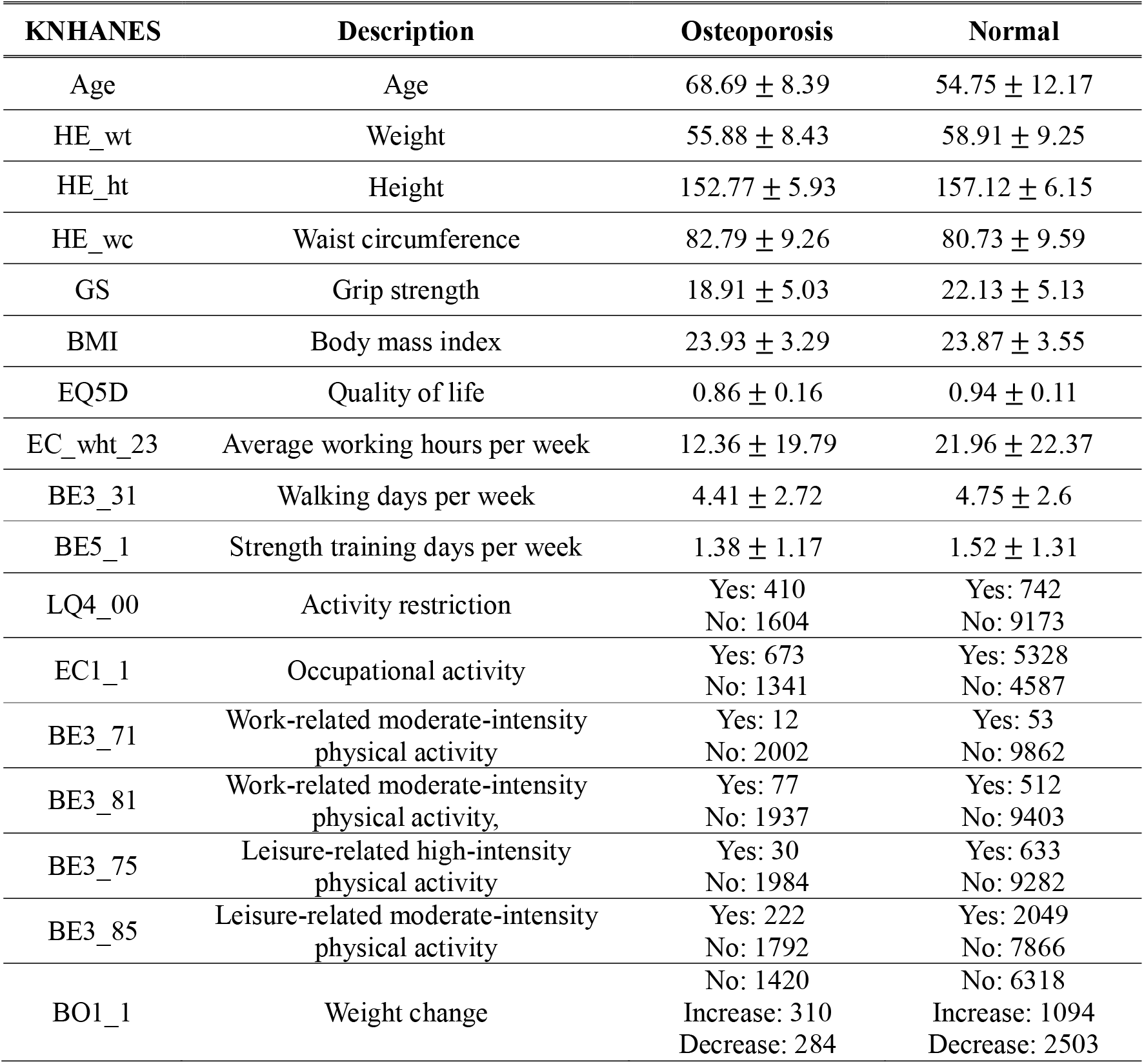
Description of Variables.

To address potential imbalances in the data between diagnosed osteoporosis (OP) and normal samples, a RandomUnderSampler function was used. This technique randomly selected an equivalent number of normal samples to match the number of OP samples. Additionally, continuous variables were scaled using the StandardScaler function, while categorical variables were transformed using one-hot encoding methods. These steps were taken to ensure that the data was prepared appropriately sfor subsequent analysis. To avoid overfitting due to many variables, LASSO rules were used to select appropriate variables before generating the multi-layer perceptron model [26]. Alpha representing the degree of regularization was set at 0.01. After the data was preprocessed, it was split into two sets: train and test data. The ratio of the train data to the test data was 7:3. This division allowed for the model to be trained on a portion of the data and then tested on a separate set to evaluate its performance.

### 3. Multi-layer Perceptron Neural Network

A multi-layer perceptron is a distributed information processing structure composed of nodes. Each layer consists of one or more nodes. The input layer receives a signal from the outside and passes the output to the hidden layer through a weighted connection. It is then computed in the hidden layer and passed to the output layer to perform computations and generate predictions. The multi-layer perceptron model created in this study consists of one input layer, one output layer, and five hidden layers. The five layers consisted of 128, 64, 32, 16, and 8 nodes, respectively. The ReLU activation function was applied to the 5 hidden layers, and the sigmoid activation function was applied to the output layer to predict the OP. To prevent overfitting, the dropout ratio was set to 0.25 and batch normalization was performed. For the gradient descent algorithm to find the minimum loss function, Adam optimizer was used. The ratio of data split and used for validation while the model was being trained was 0.2. In order to reduce the learning time for building the model, the EarlyStopping function was used to terminate learning when the validation loss value did not improve for 20 epochs.

### 4. Model evaluation

The model’s performance was assessed by generating a confusion matrix using the test data, which classified the predicted class and the actual class as TP (true positive), FP (false positive), FN (false negative), and TN (true negative). The accuracy, precision, recall, F1-score, and ROC AUC were calculated from the confusion matrix to evaluate the model’s overall performance.

## Results

### 1. Feature Selection

The variables selected using LASSO regularization were age, quality of life index, weight, grip strength, and average working hours per week.

### 2. Multi-layer Perceptron Neural Network Model Training

The multi-layer perceptron neural network model created using the selected features showed the following accuracy and loss (Figure 3). Training of the model was completed at the 39th epoch. The average accuracy of the top 5 models built in the process of model training was 77.34%.

**Figure 3.**
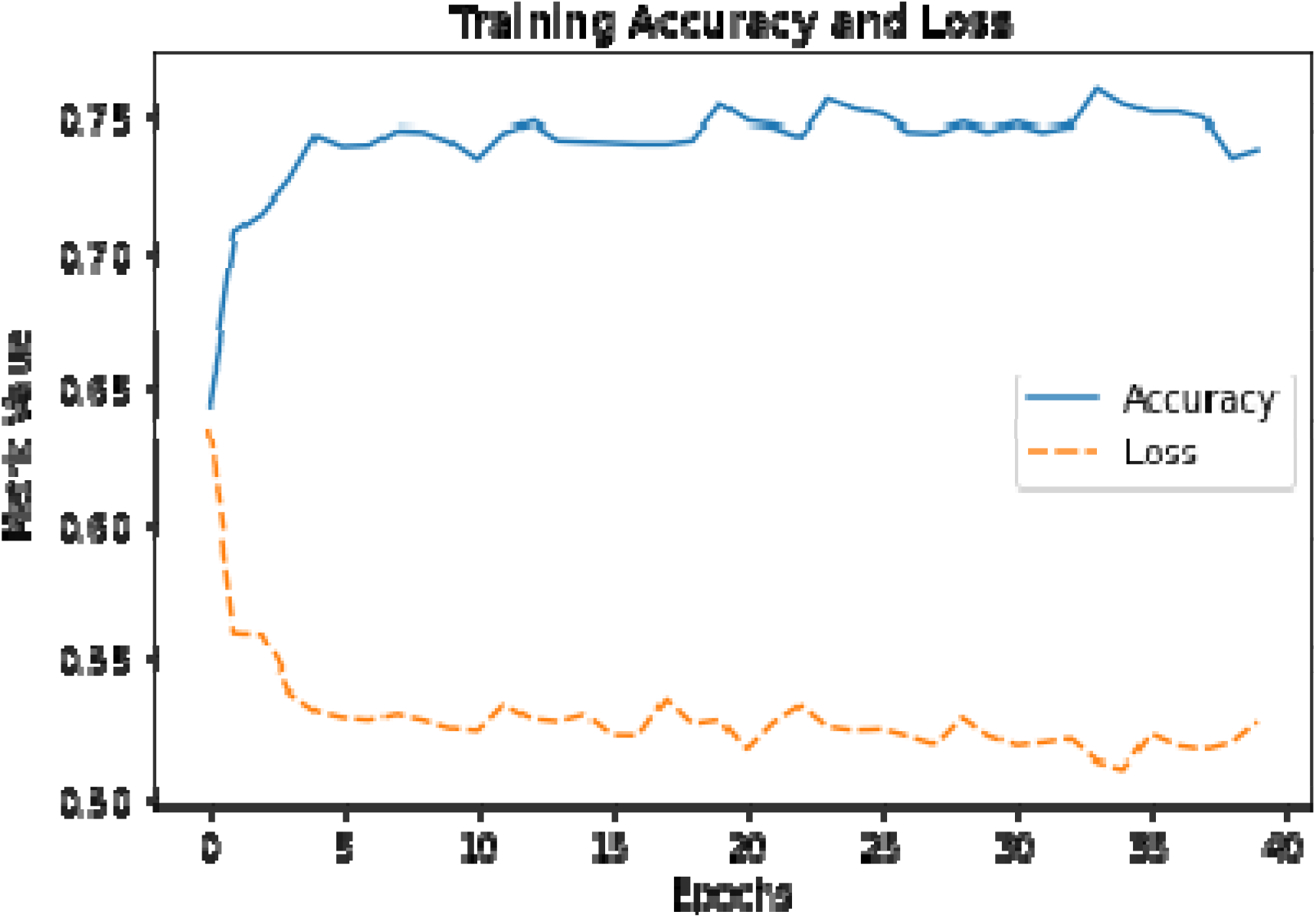
Training accuracy and loss curve.

**Figure 4.**
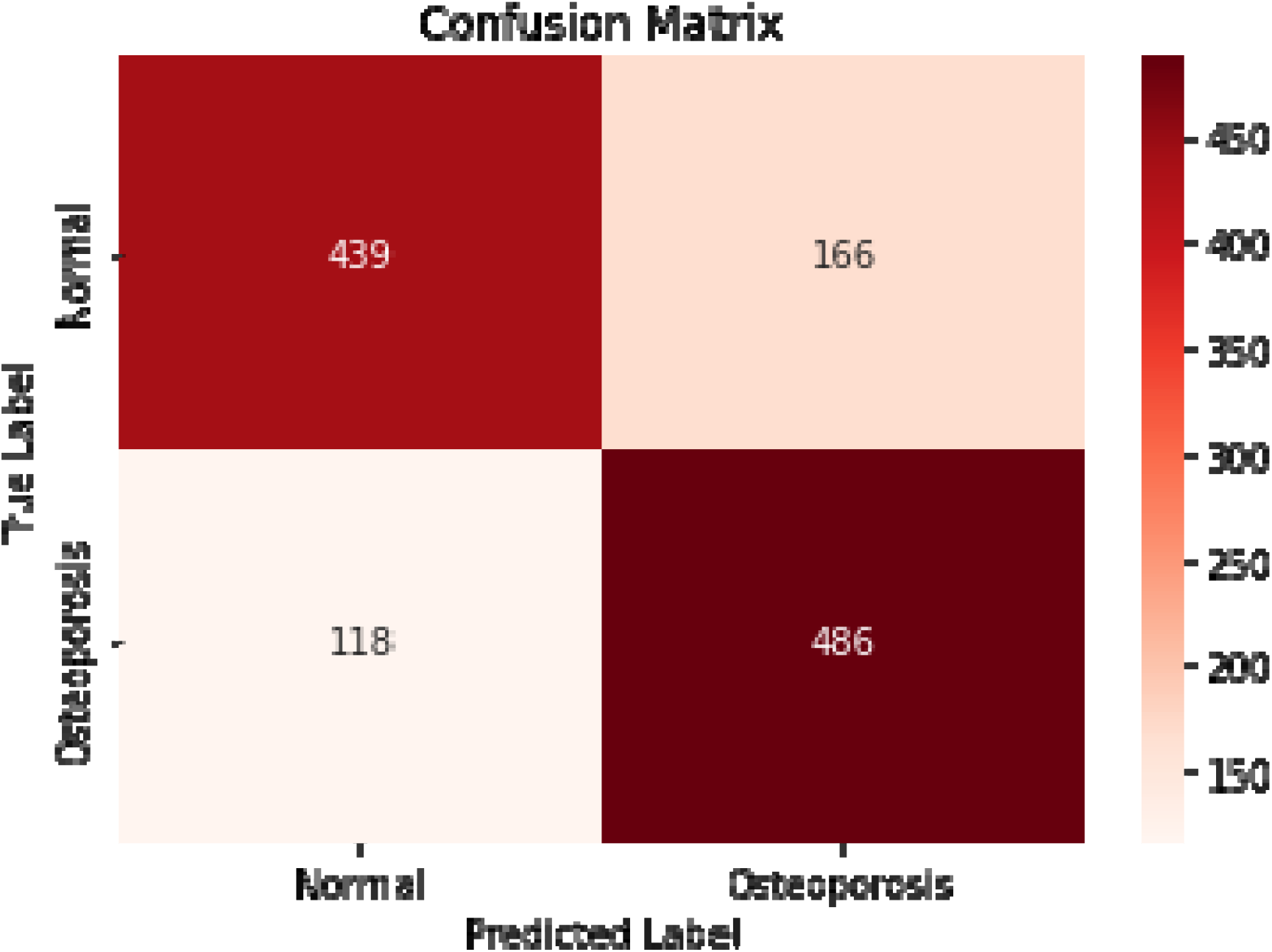
Confusion matrix for osteoporosis prediction.

### 3. Model Performance

The accuracy score, which represents the number of correctly predicted data among the entire test data by the model composed of multi-layer perceptron layer neural networks, was 76.8%. The precision score for indicating actual OP among test data samples that were predicted to have OP was 74.5%. The recall score representing the prediction of having an OP diagnosis among test data samples with an actual OP diagnosis was 80.5%. The F1 score, the harmonized mean of precision and recall, was 77.4%. The ROC AUC score, which indicates how well the multi-layer perceptron neural network model can classify each class, was 74.8%.

## Discussion

In this study, a multi-layer perceptron neural network model was created using factors related to the physical characteristics of women over the age of 35 whose bone density begins to decrease. The factors selected to create the model included not only age, weight, and grip strength, but also the quality of life and average working hours per week. All the evaluation indicators evaluating the overall performance of the model created with the above factors exceeded 74%. Among the evaluations, the recall score, which means the sensitivity of OP diagnosis, exceeded 80%.

The factors selected to model of this study may have been related to BMD, which is the criterion for diagnosis of OP. Lee et al. [27] conducted a study on the relationship between BMD and age based on the 4th KNHANES survey data between 2008 and 2009, similar to the subjects in the data sample of this study. In this study, women’s BMD was highest in the 10-19 year age group and declined at a rate of 0.66 to 1.08% per year until age 80 years [27]. Additionally, moderate physical stress can stimulate bone remodeling and increase density [28]. Therefore, factors related to physical stress, such as muscle strength and weight, appear to be related to BMD as factors that can be controlled, unlike factors such as age [28, 29]. Luo et al. [30] analyzed the relationship between grip strength and BMD based on a sample of NHANES 2013–2014 survey data of Americans, adjusting for all other factors that could affect BMD, such as age, body mass index, use of female hormones, and physical activity. Grip strength has been shown to be associated with increased femoral neck and total lumbar spine BMD in premenopausal and postmenopausal women [30]. Kim et al. [31] conducted a study on the relationship between women’s body weight and BMD.[31] In this study, they found a significant difference in weight between the abnormal group with a BMD T-score of less than -1 and the normal group with a T-score of -1 or greater. Additionally, a weak correlation existed between body weight and T-score, with each 1 kg increase in weight reducing the risk of abnormal BMD by 3.7%.[31]

The EQ-5D used to measure the quality of life variable is a tool to evaluate health-related quality of life at three levels for a total of five items: motor ability, self-management, daily life, pain and discomfort, and anxiety and depression [32]. In other words, the quality of life variable through the EQ-5D index can be seen as a variable that includes physical activity. The average working hours per week selected in the model of this study may be seen as a factor that quantitatively reflects the degree of physical activity [29, 33]. However, there is no study that directly conducted the relationship between working hours and OP, and rather, the results of other studies on the relationship between OP and work can refute this view. It has been hypothesized that endocrine disorders caused by night shifts may indirectly affect the bone physiology of night shift workers [34]. In addition, excessive physical activity may cause menstrual disorders in women, resulting in decreasing BMD [35]. Nevertheless, epidemiological evidence suggests that an active lifestyle through physical activity may reduce the incidence of osteoporotic hip fractures in the elderly population, suggesting that physical activity during working hours may be an important determinant of OP diagnosis [33, 36, 37].

Recently, models for determining OP, predicting T-score, or predicting BMD built using deep learning such as convolutional neural networks using CT and X-ray images have been proposed [38–40]. Yasaka et al. [38] built a model using 1665 CT images of 183 patients, predicted BMD values with a correlation coefficient of 0.84-0.85 with actual BMD values, and showed OP prediction performance of over 96%. Ho et al. [39] built a model with data collected through a total of 3472 pairs of pelvic X-ray and DXA examinations and predicted BMD values only with femur bone x-ray. The predicted BMD value showed a correlation r value of 0.85 with the actual value, and OP diagnosis through the model showed an accuracy of 88% [39]. In addition, Sato et al. [40] constructed a model to predict T-score with chest x-ray, which is the most common, accessible, and inexpensive measurement. The correlation coefficient between the BMD value predicted by the model and the BMD value of the hip was 0.75, and the correlation coefficient of the BMD value of the lumbar spine was 0.63 [40]. ROC AUC score, a performance indicator of the model that classified normal, osteopenia, and OP by predicting T-score, was 0.89, 0.70, and 0.84, respectively [40]. The model constructed in our study did not predict the BMD value, and the predictive performance for OP was about 75-80%, which was relatively low compared to previous studies. Nevertheless, the advantage of this model in our study is that it can predict OP with simple physical factors without imaging techniques such as CT or X-ray. The variables for using this model were easily measurable by surveying, evaluation tool, digital hand dynamometer, and did not require very high skill.

There are several limitations to this study. First, in this study, since the model was created using only female data, it is inappropriate to use male data for OP prediction. Second, it is difficult to quantitatively describe the influence of variables for OP prediction because the hidden layer of the deep learning model makes it difficult to know the exact contribution of the variables to how they arrived at the OP prediction. Therefore, it is unclear whether OP can be prevented, or symptoms improved by controlling for the variables chosen to build this model. Future studies will need to update the model with additional data or variables to improve predictive performance or to build a model for the entire population, including males. Along with this, verification of the variables selected to build this model should be performed by adjusting the influence of other extrinsic variables.

## Conclusion

In this study, a multi-layer perceptron neural network structure was created to predict the diagnosis of OP in women aged 35 years or older by using personal characteristics and physical factors as variables. The variables used to build the model were age, weight, grip strength, quality of life index, and average working hours per week, which could be easily measured through survey, assessment tool, and digital hand dynamometer. Therefore, our model will be of great help in performing OP screening without medical equipment in areas with poor medical facilities or elderly women who have difficulty accessing hospitals.

## Data Availability

All data produced are available online at Korea National Health and Nutrition Examination Survey

https://knhanes.kdca.go.kr/knhanes/sub03/sub03_02_05.do

